# Analysis of Global Ischemic Heart Disease Burden from Multiple Perspectives

**DOI:** 10.1101/2023.10.26.23297610

**Authors:** Lian Wang, Xisheng Yan, Zhengwen Xu

**Affiliations:** Department of Cardiology, Wuhan Third Hospital & Tongren Hospital of Wuhan University, Wuhan, _Hubei_, 430000, China

**Keywords:** Ischemic heart disease, Incidence, Prevalence, Death, DALYs, Age-standardised rate

## Abstract

**Background:** Ischemic heart disease (IHD) is a prevalent cardiovascular condition that poses a significant risk to human health. It has become essential to update the global prevalence, incidence, and mortality of IHD to examine secular trends.

**Methods:** The prevalence, incidence, death rates, and disability-adjusted life years (DALYs) of IHD were obtained from the Global Burden of Disease Study 2019 to assess the disease burden. We used joinpoint regression analysis to detect temporal changes and estimate the annual percent of change (APC) of each trend segment. The annual percentage change (EAPC) to access the temporal trends of the disease burden of IHD. Additionally, an analysis of the associated risk factors for IHD was conducted.

**Results:** The global number of IHD prevalence cases has increased from 96.90 million to 197.22 million, along with an increase in incidence, death, and DALYs from 1990 to 2019. After adjusting for age standardization, all indicators have shown a decrease. The decline was more pronounced among females. The number of IHD cases increased with age. In 1990 and 2019, the highest age-standardized prevalence rate (ASPR) of IHD was observed in North Africa, the Arabian Peninsula, and surrounding countries. The ASPR and age-standardized incidence rate (ASIR) of IHD were highest in low-middle SDI regions. High systolic blood pressure was the main driving force for IHD.

**Conclusions:** IHD has shown a decline in morbidity and mortality worldwide, but is on the rise in some less developed regions. The risk of incidence and death from IHD is higher in males. Paying more attention to the occurrence of IHD in the elderly is key to prevention. There exists a strong correlation between social development and the rates of morbidity and mortality associated with IHD. The inequity in global health is especially apparent in the context of this disease.

## Introduction

Ischemic heart disease (IHD) refers to a group of chronic conditions characterized by the occurrence of ischemia in the coronary arteries of the heart. This lack of blood flow leads to subsequent damage to the myocardium ^[1]^. In the year 2017, the worldwide occurrence of IHD surged to a staggering 10.6 million cases ^[2]^. The substantial impact of disease resulting from a significant population size significantly impairs individuals’ overall well-being and contributes to a heightened prevalence of severe complications, thereby intensifying the demand for cognitive resources in addressing this ailment. Epidemiological research provides compelling evidence that smoking, elevated total cholesterol levels, hypertension, obesity, and insufficient physical activity represent crucial risk factors for mortality due to IHD^[3-5]^. Over the past few decades, advancements in living conditions and healthcare accessibility have led to a reduction in the disease burden associated with IHD ^[2, 6, 7]^. Nevertheless, the substantial population size affected by IHD and the notable discrepancy in healthcare standards between the national and regional levels are significant social and public concerns that necessitate attention and resolution.

Only a few recent Global Burden of Disease (GBD) studies have comprehensively discussed IHD, and these studies have mostly focused on investigating the causes of the disease without providing a thorough analysis of its worldwide impact. Further research and comprehensive analyses are required to assist policymakers in effectively distributing and maximizing the utilization of scarce healthcare resources to mitigate the prevalence of the disease. Consequently, an initial thorough and methodical assessment of the GBD data spanning from 1990 to 2019 has been conducted. This assessment involved the classification of data based on various factors such as age, gender, SDI, region, and country. The distribution and temporal variations in the worldwide burden of illness associated with IHD from 1990 to 2019 are investigated in this study. The analysis, based on publicly accessible information, enables a comprehensive examination of risk factors contributing to DALYs and mortality, providing policymakers with more precise preventative strategies.

## Materials and methods

### Data sources

The investigation was carried out utilizing GBD data on IHD spanning the years 1990 to 2019, as available through the website of the Institute for Health Metrics and Evaluation (IHME) (https://ghdx.healthdata.org/gbd-results-tool). The GBD 2019 study represents a comprehensive revision and extension of the GBD 2017 study, using data derived from scholarly publications authored by researchers from diverse nations globally ^[8]^. The SDI, derived from the previous distribution of per capita income, the average education level of individuals aged 15 and above, and the total fertility rate among individuals under the age of 25, is utilized as an indicator to assess the comprehensive social development of a nation on a scale ranging from zero to one ^[2]^. The 204 countries and territories under consideration were categorized into distinct quintiles based on their SDI scores, namely high, high-middle, middle, low-middle, and low ^[9]^. The institutional review board of Wuhan Third Hospital & Tongren Hospital of Wuhan University, Wuhan, China, determined that the study did not need ethical approval because it used publicly available data. GBD 2019 is a publicly available database without participants’ privacy information.

### Statistical analyses

The utilization of age-standardized rates (ASR), which were determined by factoring in the weight of age distribution, serves as a dependable metric that effectively mitigates the influence of variations in age structure and population size ^[10]^. To quantify the global burden of IHD, this study used age-standardized rates (ASR) for incidence, prevalence, deaths, DALYs, and the estimated annual percentage change (EAPC) in prevalence ^[11]^, and the units of the standardized rate were per 100,000 population ^[12]^. The EAPC is used to measure the trend of ASR in a specific interval which is a summary and widely applied indicator ^[13, 14]^. The values within the EAPC possess the subsequent definition: when both the estimated value derived from the EAPC and the value at the lower boundary of its 95% confidence interval exhibit positivity, the ASR is said to have an upward trend. In contrast, when both the estimated value of the EAPC and the upper limit of its 95% confidence interval exhibit negative values, it is indicative of a declining trend in the ASR. In contrast, the ASR was deemed to be stable and unaltered according to previous studies ^[15, 16]^. The researchers employed the appropriate linear regression mathematical model to account for the natural logarithm of ASR in combination with the year ^[17]^. The joinpoint regression model was employed to determine the specific year in which significant changes occurred in the trends of the ASRs ^[18]^.

### Tools

The statistical analyses were performed using R software (R-4.3.0 version) and Joinpoint software (4.9.1 version). The visualization of the results was carried out using the ggplot2 package within the R program. Statistical significance was ascertained for two-sided p-values below 0.05.

## Result

### 1990-2019 global trends in IHD burden

From 1990 to 2019, there was a notable rise in the prevalence of cases with IHD, with the number increasing from 96.90 million (95%Uncertainty Intervals[UI]:87.04 to 107.95) to 197.22 million (95%UI:177.69 to 219.50) (Table 1). The incidence of deaths attributed to IHD exhibited an increase from 5.69 million (95%UI:5.40 to 5.9) to 9.1 million (95% UI:8.3 to 9.7) cases (Table 2). The ASPR of IHD was 2538.6 (95%UI:2282.61 to 2819.39) per 100,000 population in 1990 and 2421.02 (95%UI:2180.5 to 2692.65) per 100,000 population in 2019, with the EAPC of -0.2 (95% Confidence Interval[CI]:-0.22 to -0.18) (Table 1). The age-standardized death rate (ASDR) of IHD exhibited a drop of 136% (Table 2). The global number of IHD prevalence, incidence, death, and DALYs cases has increased. After the age standardization adjustment, all indicators have decreased. The ASPR of IHD was higher in males than in females. In 1990, the ASPR of IHD among males was estimated to be 3226.17 (95% CI: 2908.12 to 3573.10) per 100,000 population. Among females, the ASPR of IHD was estimated to be 1980.44 (95% CI: 1770.39 to 2208.72) per 100,000 population. In 2019, the ASPR of IHD among males was estimated to be 3007.47 (95% CI: 2717.43 to 3328.87) per 100,000 population. Among females, it was estimated to be 1911.53 (95% CI: 1708.95 to 2140.34) per 100,000 population.

**Table 1.** The prevalence case numbers and age-standardized prevalence of IHD in 1990 and 2019, and its temporal trends between 1990 and 2019.

| Characteristics | 1990 |  | 2019 |  | 1990-2019 |
| --- | --- | --- | --- | --- | --- |
|  | Number of prevalent cases | ASR per 100000 of prevalence | Number of prevalent cases | ASR per 100000 of prevalence | EAPC of ASPR |
|  | ×10*5 (95% UI) | (95% UI) | ×10*5 (95% UI) | (95% UI) | (95% CI) |
| Global | 968.95 ( 870.39 to 1079.53 ) | 2538.6 ( 2282.61 to 2819.39 ) | 1972.19 ( 1776.88 to 2195.01 ) | 2421.02 ( 2180.5 to 2692.65 ) | -0.2(-0.22 to -0.18) |
| Sex |  |  |  |  |  |
| Female | 412.76(368.26 to 461.09 ) | 1980.44(1770.39 to 2208.72) | 835.69(747.00 to 935.80) | 1911.53(1708.95 to 2140.34) | -0.16(-0.18 to -0.13) |
| Male | 556.18(499.74 to 619.10) | 3226.17(2908.12 to 3573.10) | 1136.51(1022.82 to 1260.35) | 3007.47(2717.43 to 3328.87) | -0.28(-0.3 to -0.26) |
| SDI |  |  |  |  |  |
| High SDI | 235.4 ( 211.23 to 260.94 ) | 2247.58 ( 2024.79 to 2486.36 ) | 314.46 ( 286.79 to 342.51 ) | 1709.47 ( 1562.14 to 1857.33 ) | -1.12(-1.2 to -1.03) |
| High-middle SDI | 283.66 ( 253.11 to 318.21 ) | 2760.77 ( 2473.53 to 3083.14 ) | 513.79 ( 458.09 to 578.36 ) | 2527.35 ( 2255.59 to 2835.5 ) | -0.38(-0.41 to -0.34) |
| Middle SDI | 238.51 ( 211.4 to 269.25 ) | 2471.34 ( 2202.2 to 2768.77 ) | 622.57 ( 553.07 to 701.93 ) | 2583.34 ( 2303.39 to 2896.24 ) | 0.18(0.16 to 0.2) |
| Low-middle SDI | 155.95 ( 140.52 to 173.52 ) | 2851.56 ( 2573.72 to 3171.25 ) | 392.03 ( 353.52 to 435.94 ) | 3010.9 ( 2721 to 3351.78 ) | 0.23(0.2 to 0.26) |
| Low SDI | 54.88 ( 49.42 to 61.01 ) | 2544.84 ( 2302.61 to 2822.23 ) | 128.19 ( 115.5 to 142.07 ) | 2659.32 ( 2402.54 to 2948.87 ) | 0.17(0.16 to 0.19) |
| Regions |  |  |  |  |  |
| High-income Asia Pacific | 24.92 ( 22.36 to 27.59 ) | 1259.52 ( 1132.58 to 1393.25 ) | 46.44 ( 41.9 to 51.22 ) | 1059.66 ( 962.51 to 1162.7 ) | -0.73(-0.82 to -0.63) |
| High-income North America | 90.42 ( 80.84 to 100.82 ) | 2549.5 ( 2281.57 to 2832.46 ) | 99.84 ( 90.2 to 109.62 ) | 1625.9 ( 1480.16 to 1776.59 ) | -1.83(-1.95 to -1.71) |
| Western Europe | 126.51 ( 112.39 to 141.11 ) | 2189.61 ( 1957.13 to 2441.13 ) | 157.57 ( 143.14 to 172.53 ) | 1810.7 ( 1650.21 to 1975.63 ) | -0.82(-0.9 to -0.73) |
| Australasia | 7.63 ( 7.07 to 8.18 ) | 3260.85 ( 3024.37 to 3498.92 ) | 13.88 ( 12.88 to 14.94 ) | 2836.79 ( 2639.76 to 3044.5 ) | -0.49(-0.52 to -0.46) |
| Eastern Europe | 96.6 ( 86.55 to 107.94 ) | 3591.54 ( 3236.58 to 3990.68 ) | 128.47 ( 115.32 to 143.35 ) | 3684.77 ( 3318.2 to 4094.89 ) | 0(-0.06 to 0.07) |
| Central Europe | 49.01 ( 43.26 to 55.91 ) | 3391.8 ( 3009.89 to 3841.41 ) | 60.06 ( 53 to 68.1 ) | 2785.63 ( 2475.39 to 3150.83 ) | -0.8(-0.84 to -0.75) |
| Southern Latin America | 6.23 ( 5.68 to 6.78 ) | 1376.51 ( 1256.31 to 1505.02 ) | 9.64 ( 8.86 to 10.45 ) | 1152.31 ( 1059.82 to 1249.58 ) | -0.64(-0.69 to -0.58) |
| East Asia | 180.76 ( 156.68 to 210.52 ) | 2181.35 ( 1901.66 to 2513.98 ) | 466.49 ( 404.27 to 540.87 ) | 2312.72 ( 2023.49 to 2660.97 ) | 0.25(0.2 to 0.3) |
| Central Asia | 17.82 ( 16.39 to 19.46 ) | 4037.68 ( 3722.72 to 4379.37 ) | 27.63 ( 25.44 to 30.06 ) | 4131.78 ( 3827.59 to 4475.15 ) | 0.02(-0.04 to 0.08) |
| North Africa and Middle East | 80.49 ( 74.58 to 87.05 ) | 5087.39 ( 4736.66 to 5477.87 ) | 199.8 ( 185.02 to 215.64 ) | 4911.06 ( 4552.67 to 5295.09 ) | -0.13(-0.16 to -0.11) |
| Southeast Asia | 40.95 ( 36.15 to 46.56 ) | 1700.39 ( 1504.12 to 1928.1 ) | 99.95 ( 88.25 to 114.17 ) | 1713.42 ( 1515.63 to 1953.3 ) | 0.01(-0.03 to 0.04) |
| Southern Sub-Saharan Africa | 5.8 ( 5.21 to 6.48 ) | 2211.12 ( 1977.3 to 2476.98 ) | 11.48 ( 10.31 to 12.79 ) | 2129.38 ( 1909.89 to 2383.49 ) | -0.25(-0.34 to -0.16) |
| Tropical Latin America | 15.17 ( 13.04 to 17.69 ) | 1726.74 ( 1481.49 to 2017.37 ) | 41.01 ( 35.33 to 47.84 ) | 1710.21 ( 1467.82 to 1995.82 ) | 0.01(-0.05 to 0.06) |
| Andean Latin America | 2.24 ( 1.95 to 2.62 ) | 1143 ( 983.94 to 1332.18 ) | 6.35 ( 5.49 to 7.41 ) | 1150.57 ( 994.68 to 1342.61 ) | 0.04(-0.02 to 0.09) |
| Caribbean | 8.61 ( 7.89 to 9.44 ) | 3335.79 ( 3055.67 to 3654.03 ) | 16.83 ( 15.42 to 18.38 ) | 3256.09 ( 2983.91 to 3554.66 ) | -0.07(-0.08 to -0.07) |
| Central Latin America | 16.46 ( 14.78 to 18.41 ) | 2064.26 ( 1838.09 to 2316.11 ) | 45.39 ( 40.57 to 51.05 ) | 1956.99 ( 1743.06 to 2196.54 ) | -0.23(-0.27 to -0.19) |
| South Asia | 167.06 ( 150.59 to 185.92 ) | 3326.82 ( 2986.31 to 3702.53 ) | 465.93 ( 420.74 to 517.26 ) | 3503.75 ( 3167.75 to 3890.14 ) | 0.21(0.17 to 0.25) |
| Central Sub-Saharan Africa | 3.71 ( 3.37 to 4.09 ) | 1805.86 ( 1639.01 to 1985.59 ) | 8.49 ( 7.75 to 9.32 ) | 1721.2 ( 1566.68 to 1887.96 ) | -0.2(-0.22 to -0.18) |
| Oceania | 0.66 ( 0.58 to 0.76 ) | 2406.97 ( 2127.29 to 2752.81 ) | 1.64 ( 1.44 to 1.89 ) | 2506.17 ( 2212.91 to 2851.2 ) | 0.15(0.11 to 0.18) |
| Western Sub-Saharan Africa | 14.86 ( 13.35 to 16.61 ) | 1782.85 ( 1602.9 to 1992.66 ) | 34.88 ( 31.26 to 39.03 ) | 1964.22 ( 1759.79 to 2202.94 ) | 0.38(0.32 to 0.44) |
| Eastern Sub-Saharan Africa | 13.02 ( 11.54 to 14.76 ) | 1846.09 ( 1641.15 to 2086.17 ) | 30.42 ( 26.98 to 34.4 ) | 1950.06 ( 1727.98 to 2210.83 ) | 0.18(0.14 to 0.22) |

IHD, Ischemic heart disease; ASR, age-standardized rate; SDI, sociodemographic index; UI, uncertainty interval; CI, confidence interval; EAPC, estimated annual percentage change;

**Table 2.** The death case number and age-standardized death of IHD in 1990 and 2019, and its temporal trends between 1990 and 2019.

| Characteristics | 1990 |  | 2019 |  | 1990-2019 |
| --- | --- | --- | --- | --- | --- |
|  | Number of death cases | ASR per 100000 of death | Number of death cases | ASR per 100000 of death | EAPC of ASDR |
|  | ×10*5 (95% UI) | (95% UI) | ×10*5 (95% UI) | (95% UI) | (95% CI) |
| Global | 56.96 ( 54.05 to 58.95 ) | 170.45 ( 159.61 to 176.94 ) | 91.38 ( 83.96 to 97.44 ) | 117.95 ( 107.83 to 125.92 ) | -1.36(-1.39 to -1.32) |
| Sex |  |  |  |  |  |
| Female | 26.73 (24.79 to 28.16 ) | 141.73(130.0 to 149.72) | 41.70 (36.80 to 45.22) | 95.07(83.91 to 103.11) | -1.50(-1.55 to -1.45) |
| Male | 30.22(28.96 to 31.33 ) | 205.24 (194.29 to 213.05 ) | 49.68 (45.91 to 53.45 ) | 144.60 (132.87 to 154.96 ) | -1.25(-1.2 to -1.22) |
| SDI |  |  |  |  |  |
| High SDI | 16.89 ( 15.73 to 17.45 ) | 162.39 ( 150.62 to 168.15 ) | 14.47 ( 12.7 to 15.54 ) | 67.1 ( 60.07 to 71.54 ) | -3.37(-3.53 to -3.21) |
| High-middle SDI | 18.71 ( 17.83 to 19.24 ) | 209.2 ( 196.25 to 216.23 ) | 26.58 ( 24.12 to 28.32 ) | 135.41 ( 122.68 to 144.44 ) | -1.73(-1.9 to -1.57) |
| Middle SDI | 11.51 ( 10.88 to 12.18 ) | 143.11 ( 133.18 to 152.13 ) | 28.25 ( 25.76 to 30.47 ) | 134.12 ( 121.51 to 145.22 ) | -0.05(-0.13 to 0.04) |
| Low-middle SDI | 7.13 ( 6.54 to 7.73 ) | 144.21 ( 132.05 to 156.58 ) | 16.46 ( 14.88 to 18.02 ) | 136.59 ( 122.96 to 149.5 ) | -0.14(-0.19 to -0.1) |
| Low SDI | 2.69 ( 2.41 to 3.02 ) | 139.2 ( 124 to 156.79 ) | 5.57 ( 4.95 to 6.27 ) | 127.99 ( 113.13 to 143.9 ) | -0.3(-0.35 to -0.24) |
| Regions |  |  |  |  |  |
| High-income Asia Pacific | 1.49 ( 1.39 to 1.55 ) | 85.48 ( 78.44 to 89.52 ) | 1.73 ( 1.4 to 1.92 ) | 30.47 ( 25.73 to 33.34 ) | -3.52(-3.73 to -3.31) |
| High-income North America | 6.55 ( 6.06 to 6.81 ) | 179.79 ( 166.53 to 186.54 ) | 6.06 ( 5.4 to 6.47 ) | 88.09 ( 79.57 to 93.4 ) | -2.85(-3.02 to -2.67) |
| Western Europe | 9.16 ( 8.6 to 9.46 ) | 156.31 ( 146.05 to 161.8 ) | 6.67 ( 5.87 to 7.18 ) | 60.69 ( 54.18 to 64.89 ) | -3.63(-3.79 to -3.46) |
| Australasia | 0.4 ( 0.38 to 0.42 ) | 178.69 ( 165.54 to 185.48 ) | 0.33 ( 0.28 to 0.36 ) | 59.04 ( 51.31 to 63.44 ) | -4.27(-4.48 to -4.07) |
| Eastern Europe | 7.96 ( 7.63 to 8.15 ) | 326.57 ( 308.96 to 336.05 ) | 9.87 ( 8.79 to 10.75 ) | 284.55 ( 253.63 to 310.14 ) | -0.92(-1.34 to -0.49) |
| Central Europe | 3.9 ( 3.72 to 3.99 ) | 300.44 ( 283.07 to 308.93 ) | 3.54 ( 3.08 to 3.95 ) | 159.8 ( 139.02 to 178.42 ) | -2.56(-2.69 to -2.44) |
| Southern Latin America | 0.66 ( 0.63 to 0.69 ) | 161.14 ( 150.08 to 167.35 ) | 0.61 ( 0.56 to 0.65 ) | 72.05 ( 65.77 to 76.66 ) | -2.75(-2.95 to -2.54) |
| East Asia | 6.31 ( 5.59 to 7.03 ) | 98.76 ( 88.2 to 109.52 ) | 19.26 ( 16.62 to 21.82 ) | 114.31 ( 98.93 to 128.75 ) | 1.13(0.86 to 1.39) |
| Central Asia | 1.32 ( 1.25 to 1.36 ) | 322.28 ( 301.66 to 333.6 ) | 2 ( 1.84 to 2.18 ) | 360.02 ( 330.9 to 390.24 ) | 0.03(-0.38 to 0.44) |
| North Africa and Middle East | 4.45 ( 4.12 to 4.78 ) | 309.32 ( 284.33 to 332.13 ) | 7.99 ( 7.06 to 9.1 ) | 219.01 ( 194.15 to 246.75 ) | -1.34(-1.42 to -1.26) |
| Southeast Asia | 7.54 ( 6.75 to 8.3 ) | 159.72 ( 143 to 177.5 ) | 18.58 ( 16.34 to 20.92 ) | 149.21 ( 130.6 to 167.88 ) | -0.24(-0.33 to -0.16) |
| Southern Sub-Saharan Africa | 0.21 ( 0.19 to 0.23 ) | 89.43 ( 80.06 to 97.89 ) | 0.44 ( 0.4 to 0.48 ) | 93.57 ( 84.43 to 102.05 ) | 0.17(-0.21 to 0.55) |
| Tropical Latin America | 1.19 ( 1.14 to 1.23 ) | 156.35 ( 145.4 to 162.47 ) | 1.76 ( 1.6 to 1.85 ) | 75.23 ( 68.22 to 79.42 ) | -2.43(-2.55 to -2.31) |
| Andean Latin America | 0.2 ( 0.17 to 0.22 ) | 109.84 ( 97.27 to 121.84 ) | 0.34 ( 0.28 to 0.41 ) | 64.48 ( 52.96 to 76.52 ) | -1.79(-2.02 to -1.55) |
| Caribbean | 0.44 ( 0.41 to 0.46 ) | 185.06 ( 172 to 193.88 ) | 0.64 ( 0.55 to 0.73 ) | 122.12 ( 106.01 to 139.69 ) | -1.51(-1.74 to -1.29) |
| Central Latin America | 0.93 ( 0.87 to 0.96 ) | 131.13 ( 121.2 to 136.69 ) | 2.19 ( 1.89 to 2.51 ) | 97.37 ( 83.84 to 111.34 ) | -1.11(-1.28 to -0.95) |
| South Asia | 7.54 ( 6.75 to 8.3 ) | 159.72 ( 143 to 177.5 ) | 18.58 ( 16.34 to 20.92 ) | 149.21 ( 130.6 to 167.88 ) | -0.24(-0.33 to -0.16) |
| Central Sub-Saharan Africa | 0.23 ( 0.19 to 0.28 ) | 130.41 ( 108.75 to 160.36 ) | 0.48 ( 0.37 to 0.62 ) | 116.78 ( 88.97 to 150.45 ) | -0.45(-0.5 to -0.4) |
| Oceania | 0.05 ( 0.04 to 0.06 ) | 183.67 ( 150.74 to 228.78 ) | 0.12 ( 0.1 to 0.15 ) | 198.93 ( 162.3 to 246.64 ) | 0.34(0.29 to 0.4) |
| Western Sub-Saharan Africa | 0.87 ( 0.72 to 1.08 ) | 126.66 ( 106.08 to 154.95 ) | 1.67 ( 1.39 to 1.95 ) | 114.61 ( 95.9 to 132.21 ) | -0.33(-0.38 to -0.28) |
| Eastern Sub-Saharan Africa | 0.59 ( 0.51 to 0.67 ) | 99.72 ( 86.19 to 111.65 ) | 1.2 ( 0.97 to 1.44 ) | 93.65 ( 74.72 to 111.98 ) | -0.26(-0.29 to -0.23) |

IHD, Ischemic heart disease; ASR, age-standardized rate; EAPC, estimated annual percentage change; UI, uncertainty interval; CI, confidence interval; SDI, sociodemographic index.

### The temporal trend in IHD burden

Globally, there has been a drop in the age-standardized incidence, prevalence, mortality, and DALYs rate of IHD from 1990 to 2019 (Figure 1). However, the line graph fails to offer a more comprehensive depiction of the fluctuations observed during the specified time frame. To be more precise, the implementation of Joinpoint analysis has allowed us to ascertain that the ASIR of IHD exhibited a decline over the periods of 1990 to 1995 (Annual percent change [APC]=0.10) and 2000 to 2005 (APC=0.15), which may be characterized as a nearly static trend. The rate of ASIR decline for IHD exhibited its most rapid pace from 2005 to 2009 (APC=1.59). Subsequently, from 2009 to 2019, the ASIR decline for IHD started to decelerate, with an APC of -0.51 (Figure 2A). The prevalence rate of the global age-standardized has experienced a notable decrease from 1990 to 2009, with the most substantial fall observed between 2001 and 2004 (APC=0.03). Nevertheless, there was a marginal incline observed from 2009 to 2014 (APC=0.03) (Figure 2B). Similarly, the joinpoint regression results of the age-standardized DALYs rate were generally similar to the findings of the age-standardized death rate, yet the magnitude of decline was larger for the age-standardized death rate. The decreasing trend of the age-standardized death rate tends to smooth out from 2017 to 2019 (APC=-0.43) (Figure 2C, Figure 2D). The disease burden associated with IHD exhibits a declining trajectory over time, but the rate of change varies at different time points.

**Figure 1.**
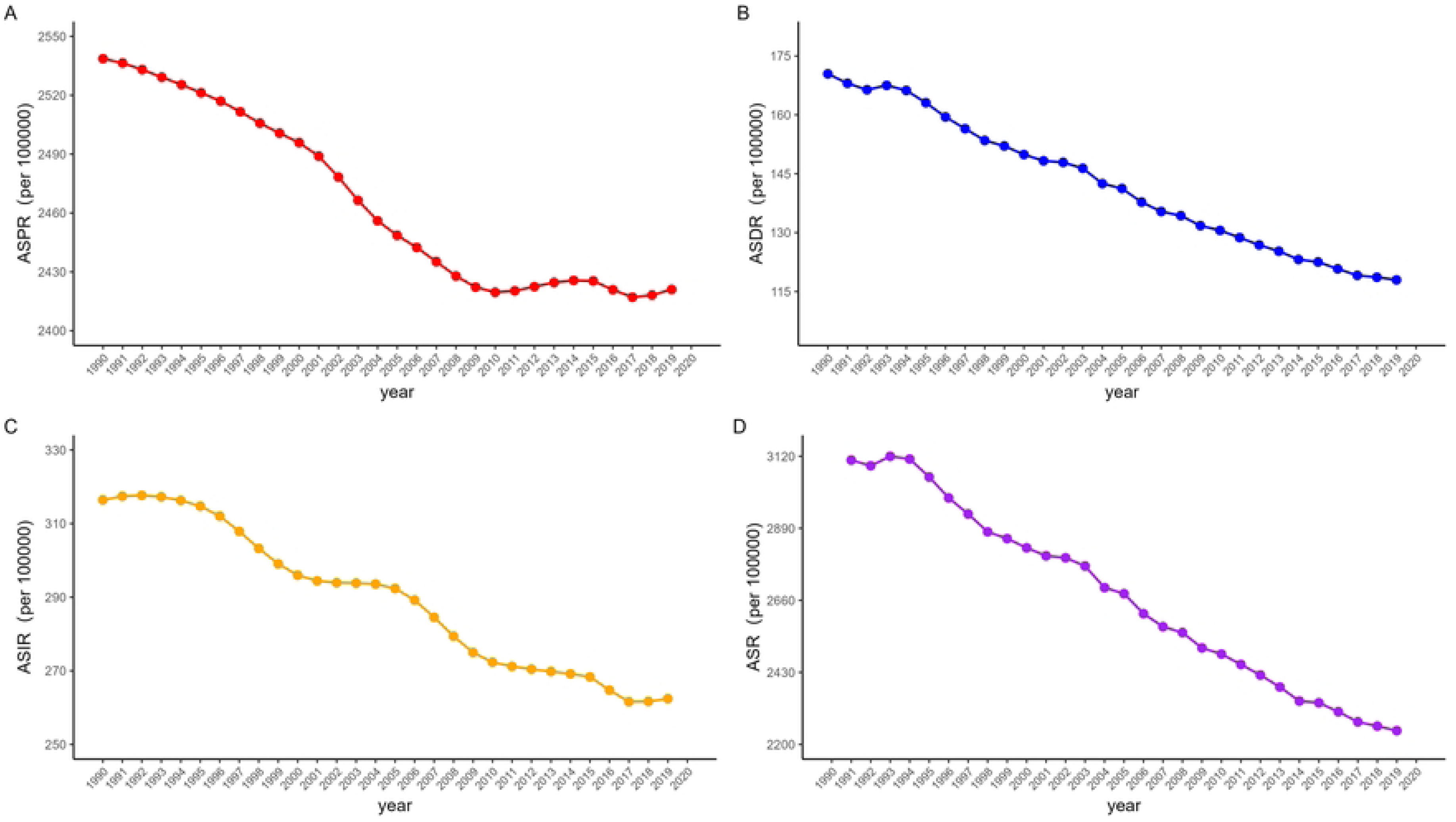

**Figure 2.**
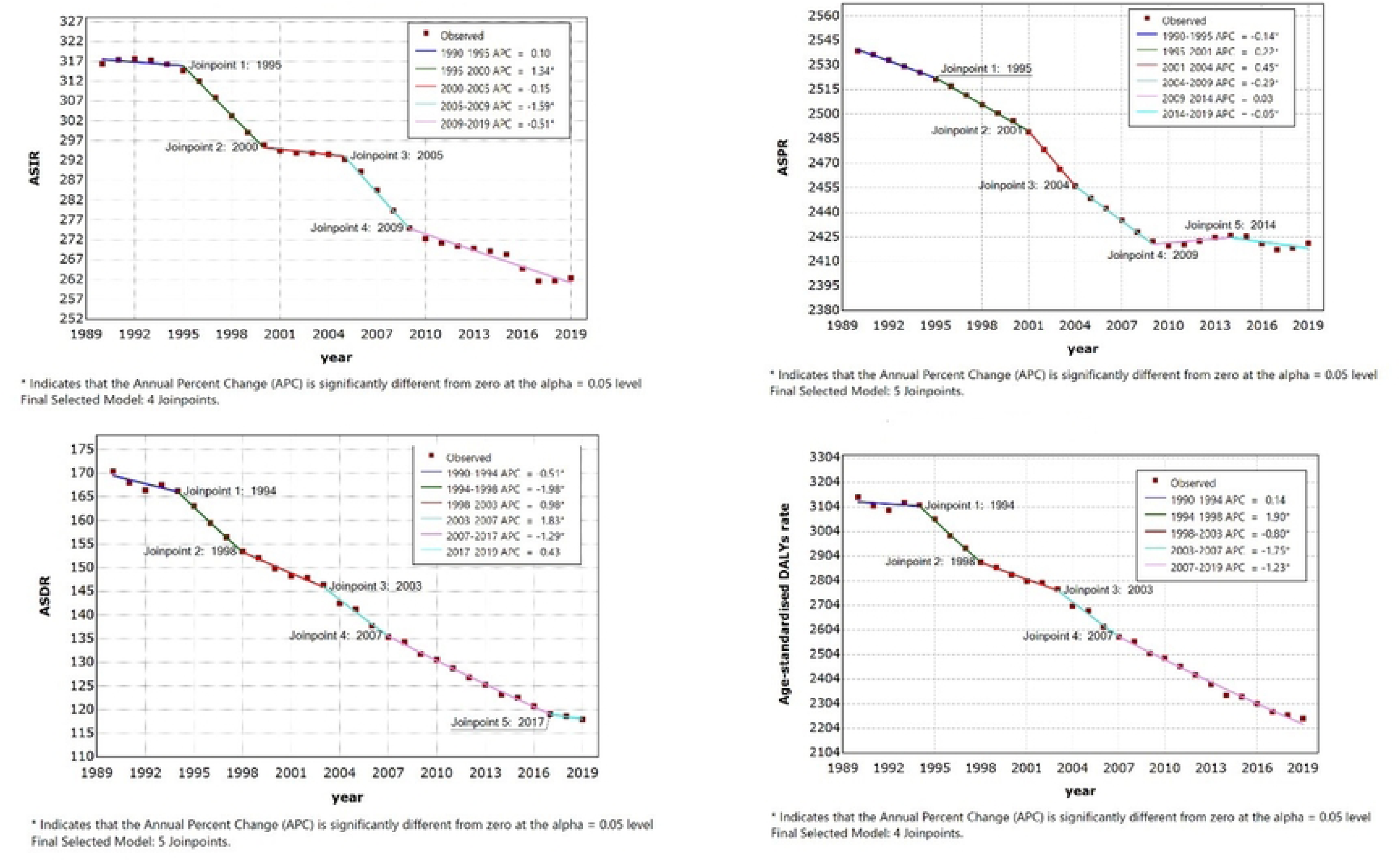

### Global distribution of IHD burden across age group and sex group

In 2019, The rates of incidence, prevalence, and DALYs exhibit an upward trend as individuals advance in age. The individuals reached their highest point of performance between the ages of 65 and 69. thereafter had a drop in performance after the age of 70 (Figure 3). The mortality rate had a positive correlation with age, and there was a notable delay of 15 years in the attainment of the peak. The incidence and prevalence of IHD and the associated DALYs were shown to be greater among females compared to males. However, it was observed that the number of deaths caused by IHD was higher among females than males specifically among individuals aged 80 years and above. The incidence of female patients with IHD was primarily observed within the age range of 65 to 84, whereas the incidence of male patients was primarily observed within the age range of 55 to 74 (Figure 4A).

**Figure 3.**
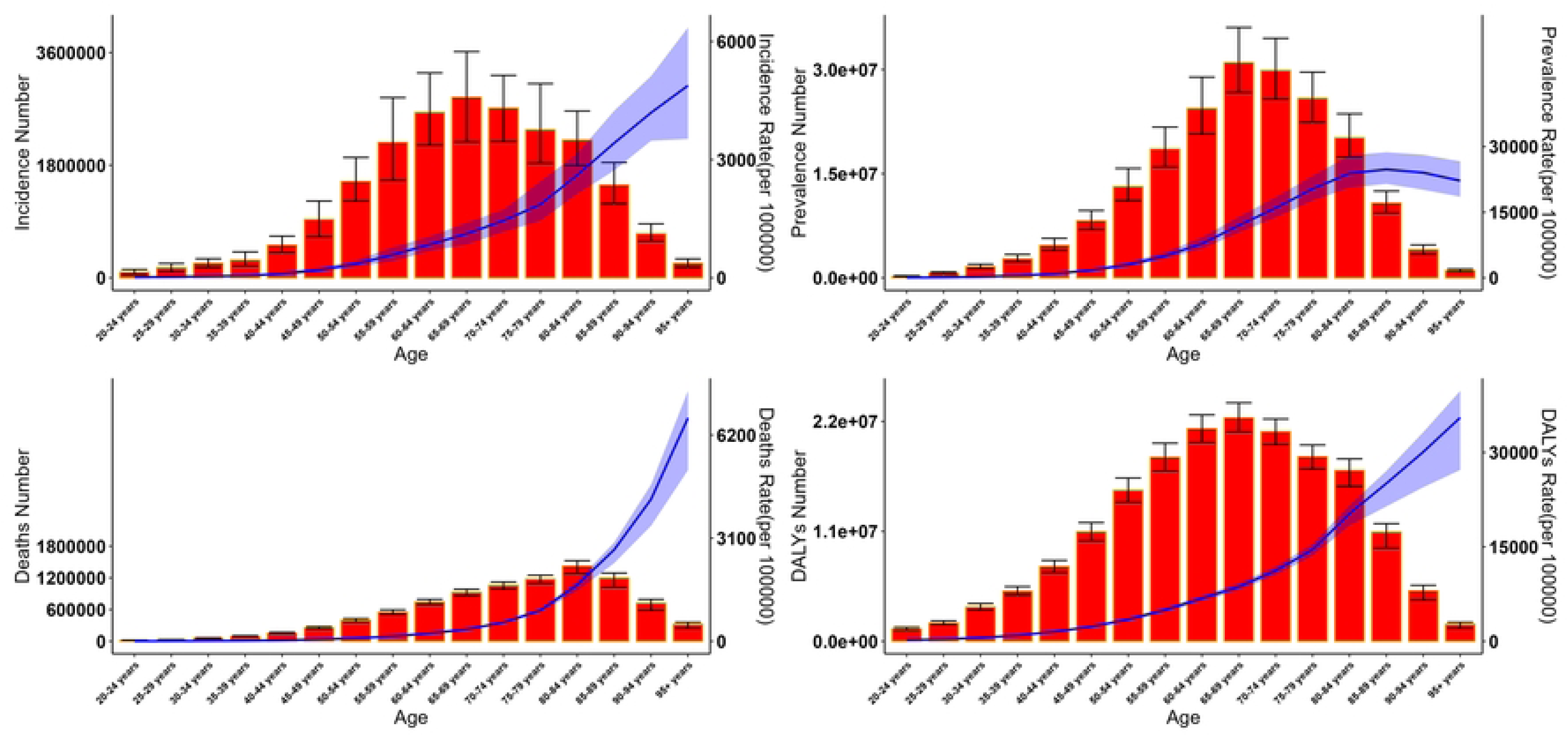

**Figure 4.**
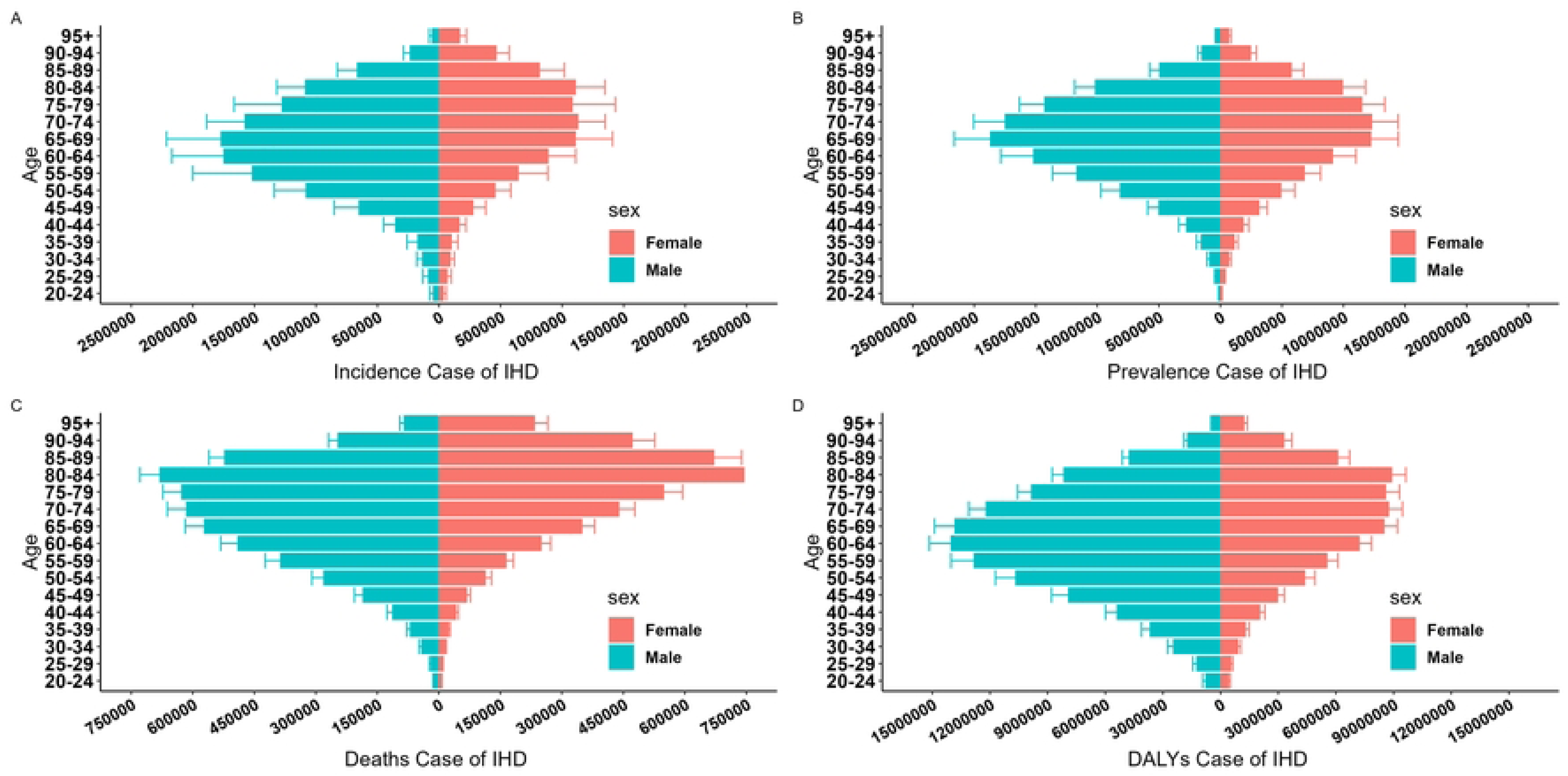

The distribution of DALYs among female patients diagnosed with IHD was mostly observed between the age range of 50 to 74. The incidence of male patients with IHD was concentrated between the ages of 60 and 84 (Figure 4D).

### Global IHD burden by region and country

In 1990, North Africa, Arabian Peninsula, and adjacent nations exhibited the highest age-standardized prevalence rates of IHD (Figure 5A). Iran’s ASPR was recorded as 6250.64 (95%UI:6852.20 to 5678.67) per 100,000 population. Following closely behind was Iraq, with a rate of 5603.59 (95% UI:5193.04 to 6053.42) per 100,000 population. The regions of Western South America and certain countries in East and Southeast Asia exhibited the lowest ASPR of IHD (Figure 5A). The countries exhibiting the lowest ASPR were Peru, with a rate of 1102.08 (95%UI:952.46 to 1284.12) per 100,000 population, and the Republic of Korea, with a rate of 1103.58 (95% UI:1007.01 to 1200.38) per 100,000 population. After a span of thirty years characterized by various transformations, it was observed that in 2019, the regions with the highest and lowest ASPR have remained unaltered and continue to be concentrated in the aforementioned locations (Figure 5B). Additionally, we analyzed the alteration in ASPR across 204 countries. The nations that exhibited the highest shift in prevalence, as measured by the EAPC, were Uzbekistan (0.86), Guinea (0.65), Chad (0.59), United Republic of Tanzania (0.54), and Guatemala (0.50), the ASPR of IHD has exhibited varied degrees of growth during the period from 1990 to 2019 (Figure 6A). From 1990 to 2019, by country and region, the top five countries with negative global prevalence growth rates were the United States (-1.90), Finland(-1.26), Poland (-1.21), Canada(-1.16), Denmark (-1.15) (Figure 6B). From the aforementioned, it becomes evident that there exist notable disparities in the epidemiological indicators of IHD across different countries.

**Figure 5.**
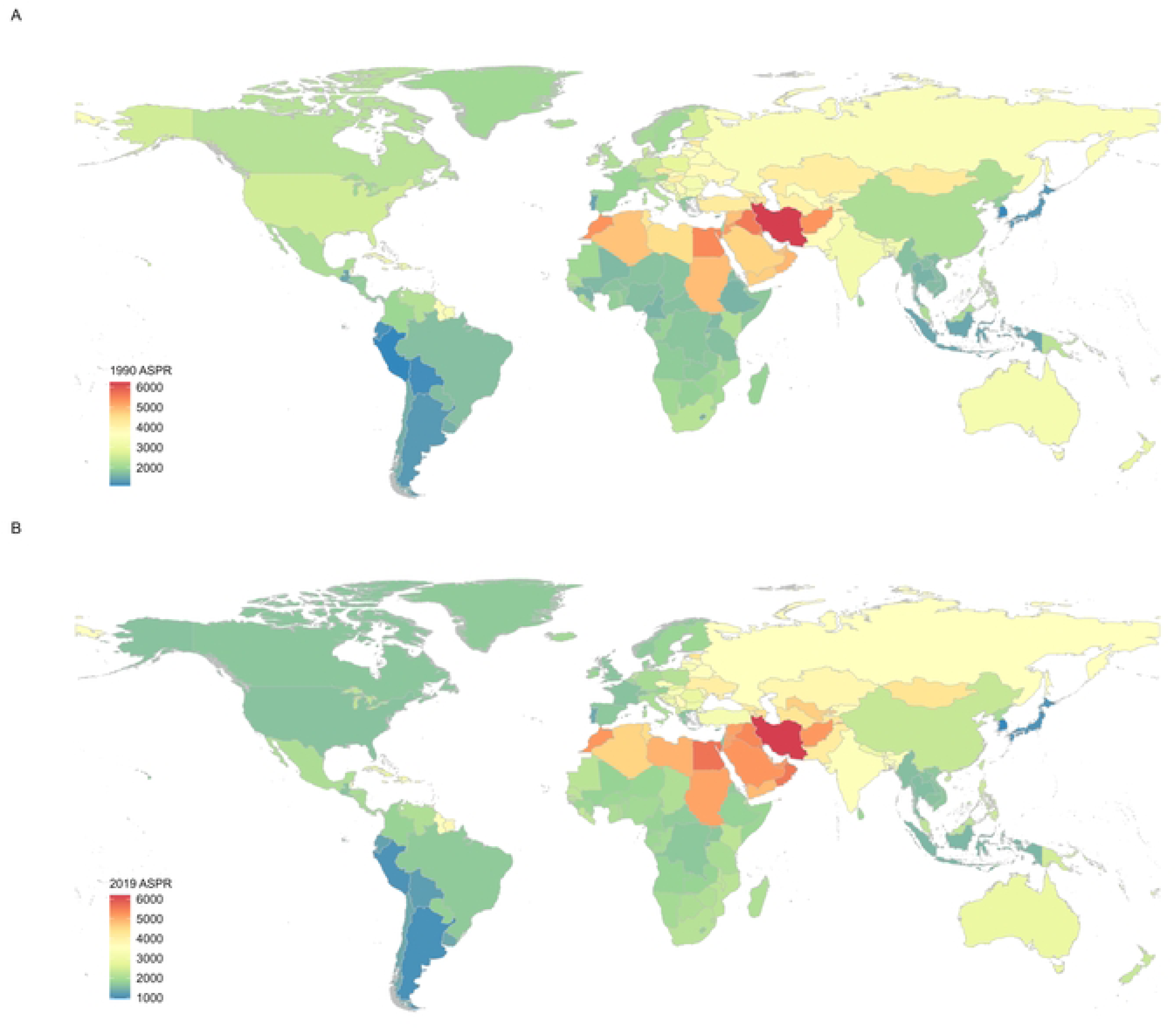

**Figure 6.**
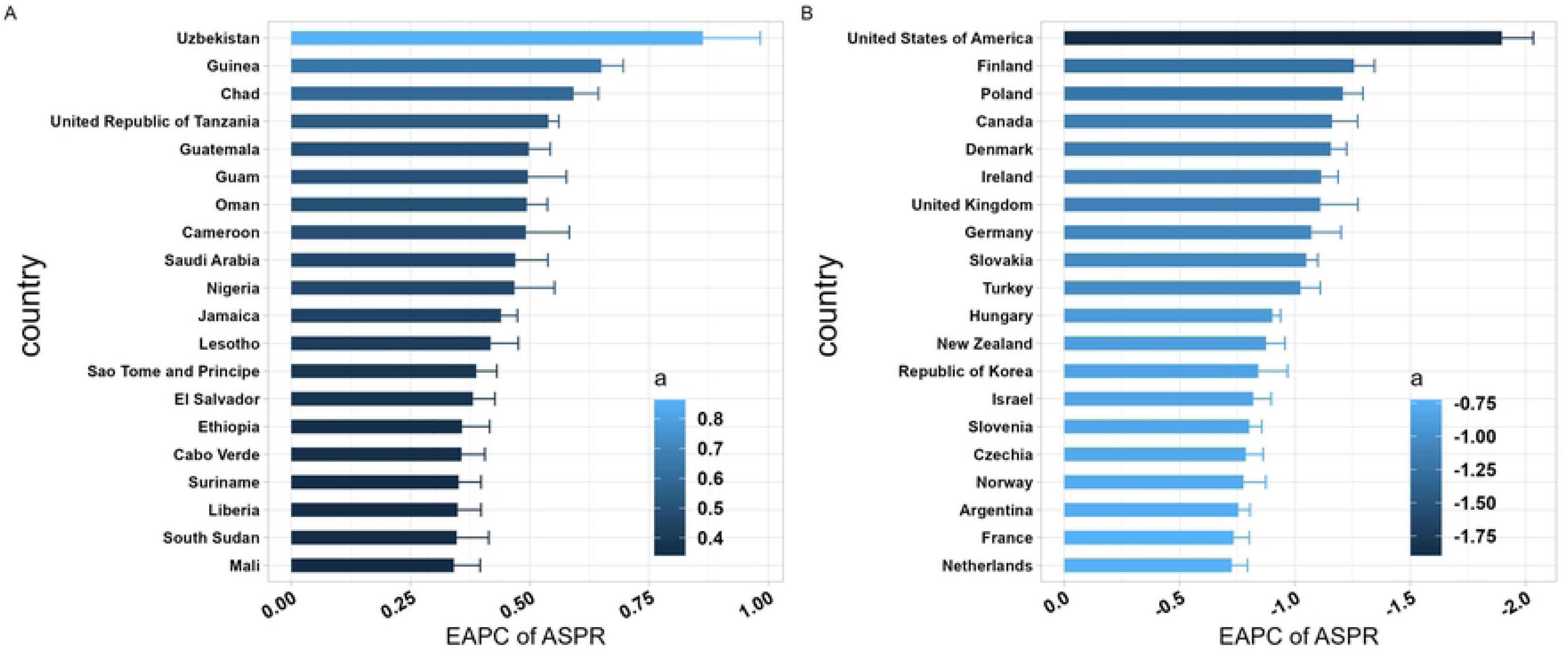

### Relation between IHD burden and SDI

From 1990 to 2019, the burden of ischemic heart disease varied considerably in levels and trends across the five SDI regions. The ASPRs and ASIRs of IHD have the highest values in regions characterized by low and medium SDI levels, while they were comparatively lower in regions with high SDI levels. The data presented in Figures 7C and D demonstrates that there was variation in ASDRs and DALYs among countries and areas with different levels of SDI. Specifically, the medium and high SDI regions exhibit the greatest rates of ASDRs and DALYs (Figure 7). Ischemic heart disease-related ASIR by SDI for both sexes combined, between 1990 and 2019 (Figure 8). The ASIR of ischemic heart disease with SDI less than 0.5 interval shows a decreasing trend. Moreover, the ASIR for IHD demonstrates an upward trajectory within the SDI range of 0.65 to 0.72, particularly in Eastern Europe. It peaked at approximately 0.72 of SDI and then showed a decreasing trend. In the relationship between SDI and ASIR in different countries we can see that the value of SDI and ASIR show a positive trend before the value of SDI reaches 0.75, and then the higher the SDI the smaller the value of ASIR (Figure 9).

**Figure 7.**
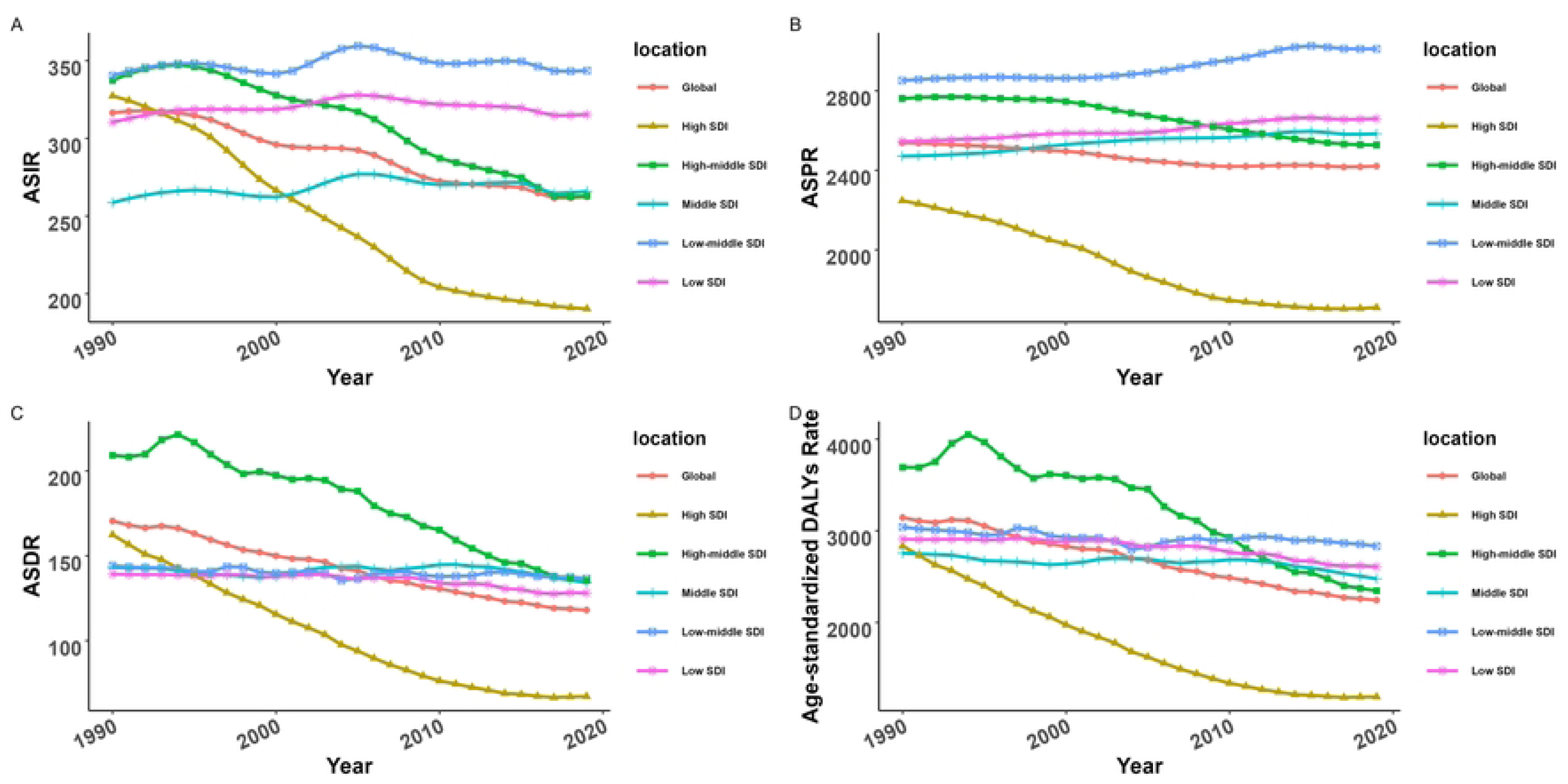

**Figure 8.**
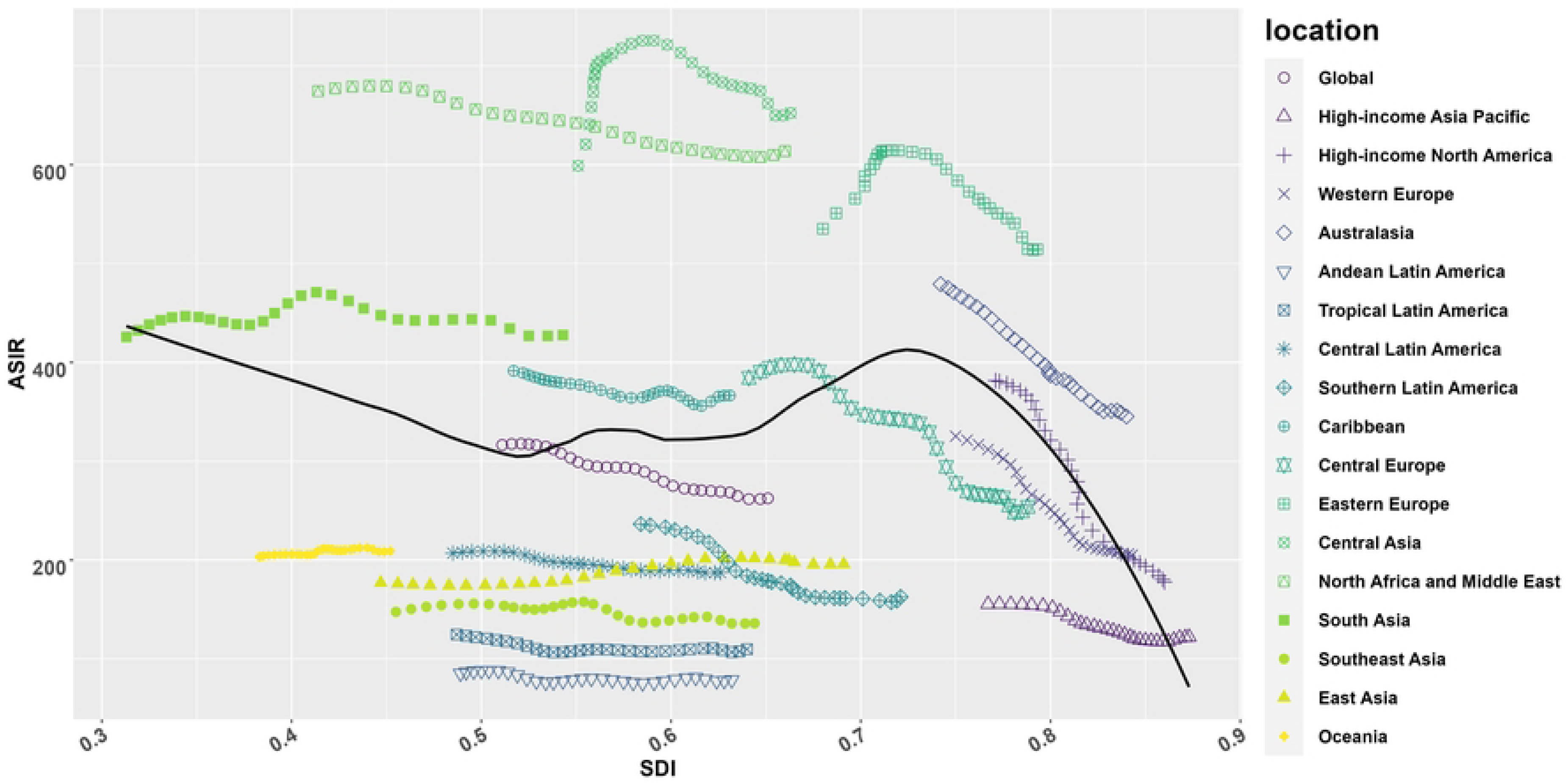

**Figure 9.**
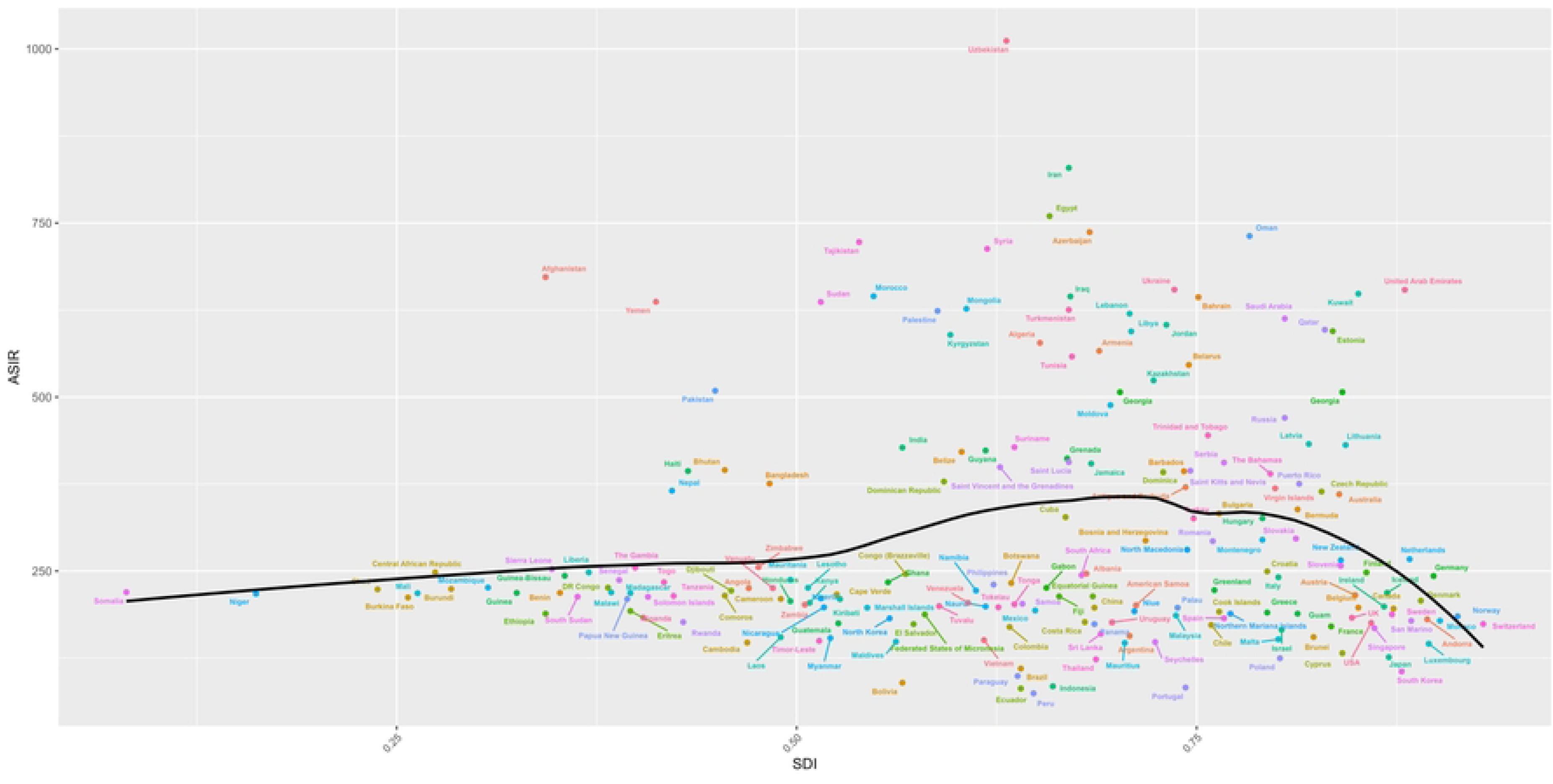

### The attributable burden of IHD to risk factors

We searched the GBD database to identify prospective risk variables associated with ischemic heart disease-related mortality and disability-adjusted life years (DALYs).In 2019, the leading risk factors for deaths and DALYs due to IHD were high systolic blood pressure(DALYs:54.3%; Deaths:52.8%), high Low-Density Lipoprotein (LDL) cholesterol (DALYs:46.2%; Deaths:41.1%), etc. High SDI regions have the lowest percentage of DALYs and deaths due to high systolic blood pressure. Based on the 21 regions analyzed in the GBD study, the percentage of DALYs and deaths attributable to high systolic blood pressure was highest in Southern Sub-Saharan Africa, and lowest in Oceania (Deaths:42.3 %) and Andean Latin America(DALYs:43.1%) (Figure 10B). The ranking of the primary risk factors contributing to mortality and disability-adjusted years caused by IHD has exhibited no change over the past three decades. Notably, high systolic blood pressure remains the foremost risk factor in terms of its prevalence among all identified risk variables. The global risk factors for deaths and disability-adjusted life years (DALYs) attributed to IHD exhibited a decline of 1.4 and 0.1, respectively. Furthermore, a significant reduction in the proportion of all risk factors was observed across numerous regions (Figure 10A, B).

**Figure 10A.**
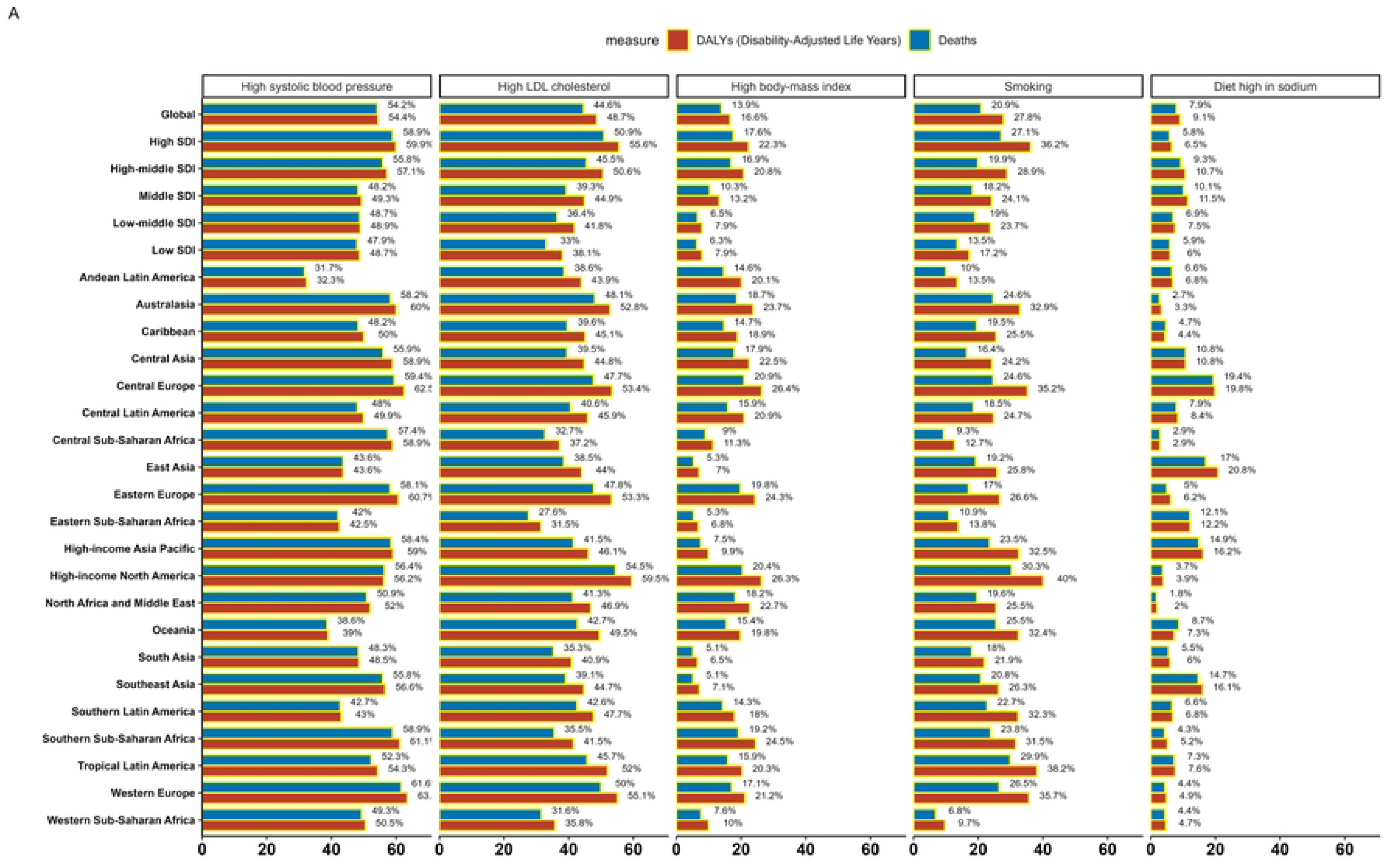

**Figure 10B.**
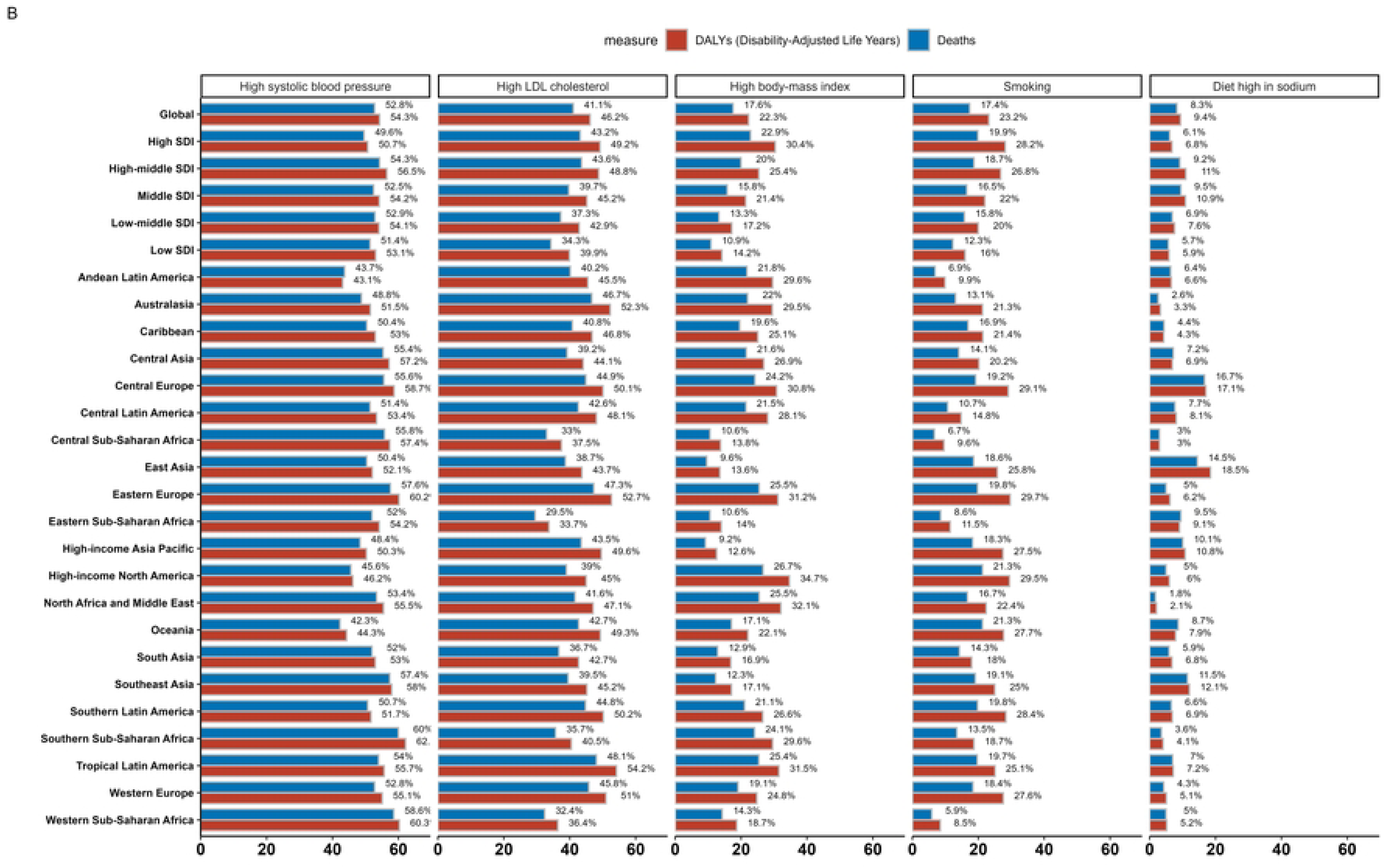

## Discussion

This project aims to assess the global spatial and temporal patterns of IHD across 204 nations and territories worldwide from 1990 to 2019. The intention is that the findings will inform strategies for enhancing the treatment of IHD. The worldwide disease burden of IHD is likely to be significantly more severe than what is commonly perceived. In 1990, the worldwide prevalence was recorded at 96 million individuals. In 2019, the prevalence of individuals affected by IHD had escalated by an additional 100 million. However, the prevalence rate showed a decline following adjustment for age standardization. Additionally, the ASDR has reached a point of stability, with the previous notable fall in recent years no longer observed. The prevalence cases and incidence cases are rising, potentially due to advancements in life expectancy and the growth of the global population.

The analysis of gender and age groups reveals a higher prevalence of IHD within the male population as compared to the female population. The increased prevalence and mortality of IHD in males relative to females can be attributed, in part, to the regulatory function of estrogen in the cardiovascular system mediated by the estrogen receptor ^[19, 20]^. According to previous studies, the weighty social obligations and significant mental and psychological stress experienced by males render them vulnerable to adopting harmful lifestyle practices, like staying up late, smoking, consuming excessive amounts of alcohol, and adhering to high-fat diets ^[21, 22]^. To prevent IHD disease, a thorough examination of physical and psychological factors is required, with a particular focus on the male susceptibility to IHD. Research shows the typical woman with IHD is old and frail, with comorbidities such as renal failure confirmed in our study ^[23]^. And as estrogen levels decline, females are exposed to the same cardiovascular risks as males. Postmenopausal females demonstrate a greater burden of cardiovascular risk factors, which, together with microvascular dysfunction and narrower and stiffer arteries, contribute to the poorer prognosis observed in females with IHD.^[24]^. Advancements in the field of medicine, coupled with a growing focus on self-care practices in maintaining one’s health, have played a significant role in the observed rise in life expectancy. In comparison to three decades ago, the age at which individuals typically experience the onset of age-related health issues has been postponed by around 25 years. Nevertheless, there was a significant increase in ASRs of IHD among individuals aged 50 years and above in both 1990 and 2019. The analysis of both the age distribution and the evolving characteristics of the burden of IHD indicates the necessity of directing increased attention towards the occurrence of IHD in the elderly population. Cardiovascular disease is widely recognized as the primary cause of mortality and significant impairment among those aged 75 years and above ^[25]^. Consequently, it is imperative, particularly in nations experiencing a rise in elderly populations, to prioritize the implementation of efficient preventive measures and optimal treatment strategies for IHD. This necessitates increased attention from elderly individuals themselves, governmental bodies, and healthcare professionals.

There has been a notable decline in the occurrence of IHD in certain Western and economically advanced nations throughout the past thirty years. Conversely, economically disadvantaged countries, including Uzbekistan, Guinea, Chad, and the United Republic of Tanzania, have had an upward trajectory in IHD incidence. IHD exhibits a higher prevalence among patients residing in low-income nations, leading to a comparatively elevated 1-year mortality rate in comparison to other disorders ^[26]^. Countries with high SDI have performed impeccably in all aspects of IHD disease control. This may reflect the fact that developed regions have higher levels of resource allocation, a more robust healthcare system, and relatively better educated patients who are more motivated to treat and prevent IHD and its complications ^[27, 28]^, which are critical to reducing the burden of disease associated with IHD. The inverse independent association between socioeconomic status and the risk of IHD in the general population has been well documented ^[29, 30]^. Global health inequalities need to be further explored, with a focus on IHD morbidity and mortality in relatively less economically developed regions and countries. Countries at disparate socio-economic levels encounter diverse risk factors and ought to be specifically addressed based on the distinct attributes of each region, to formulate suitable measures for prevention and control.

Interestingly some medium SDI countries show a higher incidence of IHD than low SDI countries, concentrated in the Middle East and North Africa (MENA) region. Studies indicate that arid climates have serious and potentially life-threatening effects on human health, particularly for people over the age of 65 ^[31]^. High incidence and mortality rates are also attributable to artificially low case diagnosis rates and misclassification of potential causes of death, as well as inadequate healthcare systems and a shortage of cardiac specialists ^[32]^. Therefore, we not only should consider the disease itself but also some factors other than the disease, perhaps these factors can also cause or aggravate the occurrence of the disease.

High systolic blood pressure continues to be the foremost risk factor for mortality and DALYs associated with IHD. The decrease of key risk factors for IHD has been achieved by advancements in medicine and the development of diverse antihypertensive medications. Numerous prior research has demonstrated that implementing rigorous blood pressure control yields improvements in left ventricular ejection fraction and the reversal of cardiac remodeling ^[33]^. The study aims to investigate the potential impact of increasing hypertension medication on the occurrence of IHD. Modifying the incidence of IHD by intensifying the treatment of hypertension ^[34]^. The treatment of blood lipids is of utmost importance, as research has demonstrated a favorable association between elevated levels of LDL and the incidence of IHD ^[35]^. The aforementioned risk factors can be effectively managed by the use of pharmaceutical interventions. Nevertheless, the persistently high death load can be attributed to factors such as the substantial population size and the diverse economic circumstances across different nations. Drawing upon the preceding analysis the prevalence of IHD is influenced by social and economic development, either directly or indirectly. Furthermore, these findings shed light on the existence of global health disparities and inequalities.

### Limitation

This is the first and most comprehensive epidemiological study to analyze the burden of IHD at the global, regional, and national levels according to SDI, sex, and age, and elucidate its trend from 1990 to 2019. However, it must be acknowledged that there are some limitations. Firstly, due to the limitation of diagnostic accuracy, underreporting and misdiagnosis may cause deviations in the true number of incident cases, incidence rates, and corresponding age-standardized rates, especially in some underdeveloped countries. Secondly, the article solely focuses on elucidating the epidemiological characteristics of the disease, without delving into a thorough exploration of its etiology, and the profound exploration of the influence exerted by the environment on IHD has been somewhat limited. Thirdly, previous studies have shown that IHD is affected by COVID-19, but due to the impact of database updates, this factor cannot be included at present. Finally, as this study was an analysis of the available data from GBD 2019, we have no detailed data to further control bias from other important risk factors for IHD including lifestyle, occupation, ethnicity, and air pollution.

### Conclusions

The global incidence and mortality rates of ischaemic heart disease have seen a downward trend. However, certain underdeveloped regions have experienced an increase. Furthermore, the risk of developing the condition and experiencing fatal outcomes is disproportionately higher among males compared to females. The age of onset is concentrated between 55 and 79 years, while the age of mortality is primarily concentrated between 75 and 89 years. The distribution of age and the evolving nature of the burden of IHD indicate the need for increased focus on preventing IHD in the elderly. There is a notable correlation between economic challenges and the morbidity and mortality associated with ischaemic heart disease. The inequity in global health is especially apparent in the context of this disease.

#### Abbreviations

GBD: Global burden of disease
IHD: Ischemic heart disease
IHME: Institute for health metrics and evaluation
SDI: Socio-demographic index
ASR: Age-standardized rate
ASPR: Age-standardized prevalence rate
ASDR: Age-standardized death rate
DALYs: Disability adjusted life years
APC: Annual percent change
EAPC: Estimated annual percentage change
UI: Uncertainty intervals
CI: Confidence interval

## Data Availability

All relevant data are within the manuscript and its Supporting Information files.

https://ghdx.healthdata.org/gbd-results-tool.

## Acknowledgments

We acknowledge The Global Burden of Disease, Injuries, and Risk Study (GBD) 2019, which presented detailed information.

## Ethics Statement

The institutional review board of Wuhan Third Hospital & Tongren Hospital of Wuhan University, Wuhan, China, determined that the study did not need ethical approval because it used publicly available data. GBD 2019 is a publicly available database without participants’ privacy information.

## Patient consent for publication

Not applicable.

## Authors’ contributions

Conceptualization: WL and XZ conceived and designed the study. Methodology: WL developed or contributed to the methodology.Writing - Original Draft Preparation: WL wrote the initial draft of the manuscript. Writing - Review and Editing: YX reviewed and edited the manuscript.Visualization: WL created visualizations or figures. Supervision: XZ provided supervision or oversight for the research project.

## Availability of data and materials

Publicly available datasets were analyzed in this study. The data can be found here: https://ghdx.healthdata.org/gbd-results-tool. Data availability statement all data relevant to the analysis are available online on their respective websites.

## References

[1] Roth G A, Mensah G A, Johnson C O, et al. Global Burden of Cardiovascular Diseases and Risk Factors, 1990-2019: Update From the GBD 2019 Study. Journal of the american college of cardiology, 2020, 76(25): 2982–3021.

[2] Wang F, Yu Y, Mubarik S, et al. Global Burden of Ischemic Heart Disease and Attributable Risk Factors, 1990-2017: A Secondary Analysis Based on the Global Burden of Disease Study 2017. Clinical epidemiology, 2021, 13(null): 859–70.

[3] Khalili D, Sheikholeslami F H, Bakhtiyari M, et al. The incidence of coronary heart disease and the population attributable fraction of its risk factors in Tehran: a 10-year population-based cohort study. PloS one, 2014, 9(8): e105804.

[4] Miller M, Zhan M, Havas S. High attributable risk of elevated C-reactive protein level to conventional coronary heart disease risk factors: the Third National Health and Nutrition Examination Survey. Archives of internal medicine, 2005, 165(18): 2063–8.

[5] Young F, Capewell S, Ford E S, et al. Coronary mortality declines in the U.S. between 1980 and 2000 quantifying the contributions from primary and secondary prevention. American journal of preventive medicine, 2010, 39(3): 228–34.

[6] Khan M A, Hashim M J, Mustafa H, et al. Global Epidemiology of Ischemic Heart Disease: Results from the Global Burden of Disease Study. Curēus, 2020, 12(7): e9349.

[7] Wang L, Ma N, Wei L. Global burden of ischemic heart disease attributable to high sugar-sweetened beverages intake from 1990 to 2019. Nutrition metabolism and cardiovascular diseases, 2023, 33(6): 1190–6.

[8] Global burden of 369 diseases and injuries in 204 countries and territories, 1990- 2019: a systematic analysis for the Global Burden of Disease Study 2019. Lancet, 2020, 396(10258): 1204–22.

[9] Du M, Chen W, Liu K, et al. The Global Burden of Leukemia and Its Attributable Factors in 204 Countries and Territories: Findings from the Global Burden of Disease 2019 Study and Projections to 2030. Journal of Oncology, 2022, 2022(null): 1612702.

[10] Jin Z, Wang D, Zhang H, et al. Incidence trend of five common musculoskeletal disorders from 1990 to 2017 at the global, regional and national level: results from the global burden of disease study 2017. Annals of the rheumatic diseases, 2020, 79(8): 1014–22.

[11] Ding Q, Liu S, Yao Y, et al. Global, Regional, and National Burden of Ischemic Stroke, 1990-2019. Neurology, 2022, 98(3): e279–e90.

[12] Li H, Song X, Liang Y, et al. Global, regional, and national burden of disease study of atrial fibrillation/flutter, 1990-2019: results from a global burden of disease study, 2019 [J]. BMC public health, 2022, 22(1): 2015.

[13] Smith-Bindman R, Kwan M L, Marlow E C, et al. Trends in Use of Medical Imaging in US Health Care Systems and in Ontario, Canada, 2000-2016. Jama-journal of the american medical association, 2019, 322(9): 843–56.

[14] Qin Y, Tong X, Fan J, et al. Global Burden and Trends in Incidence, Mortality, and Disability of Stomach Cancer From 1990 to 2017. Clinical and translational gastroenterology, 2021, 12(10): e00406.

[15] Hu Y, Tong Z, Huang X, et al. The projections of global and regional rheumatic heart disease burden from 2020 to 2030. Frontiers in cardiovascular medicine, 2022, 9(null): 941917.

[16] Liu Z, Jiang Y, Yuan H, et al. The trends in incidence of primary liver cancer caused by specific etiologies: Results from the Global Burden of Disease Study 2016 and implications for liver cancer prevention. Journal of hepatology, 2019, 70(4): 674–83.

[17] Hankey B F, Ries L A, Kosary C L, et al. Partitioning linear trends in age-adjusted rates. Cancer causes & control, 2000, 11(1): 31–5.

[18] Jung K W, Won Y J, Hong S, et al. Prediction of Cancer Incidence and Mortality in Korea, 2020. Cancer Research and Treatment, 2020, 52(2): 351–8.

[19] Mishra S R, Chung H F, Waller M, et al. Duration of estrogen exposure during reproductive years, age at menarche and age at menopause, and risk of cardiovascular disease events, all-cause and cardiovascular mortality: a systematic review and meta-analysis. Bjog-an international journal of obstetrics and gynaecology, 2021, 128(5): 809–21.

[20] Xiang D, Liu Y, Zhou S, et al. Protective Effects of Estrogen on Cardiovascular Disease Mediated by Oxidative Stress. Oxidative Medicine and Cellular Longevity, 2021, 2021(null): 5523516.

[21] Li Y, Zhang J. Disease burden and risk factors of ischemic heart disease in China during 1990-2019 based on the Global Burden of Disease 2019 report: A systematic analysis. Frontiers in public health, 2022, 10(null): 973317.

[22] Papier K, Knuppel A, Syam N, et al. Meat consumption and risk of ischemic heart disease: A systematic review and meta-analysis. Critical reviews in food science and nutrition, 2023, 63(3): 426–37.

[23] Andreotti F, Rio T, Gianmarinaro M, et al. [Pathophysiology of ischemic heart disease in women. Giornale Italiano di Cardiologia, 2012, 13(6): 396–400.

[24] Dorobantu M, Onciul S, Tautu O F, et al. Hypertension and Ischemic Heart Disease in Women. Current pharmaceutical design, 2016, 22(25): 3885–92.

[25] Amsterdam E A, Wenger N K, Brindis R G, et al. 2014 AHA/ACC Guideline for the Management of Patients with Non-ST-Elevation Acute Coronary Syndromes: a report of the American College of Cardiology/American Heart Association Task Force on Practice Guidelines. Journal of the american college of cardiology, 2014, 64(24): e139–e228.

[26] Tromp J, Bamadhaj S, Cleland J, et al. Ischemic heart disease is more prevalent in low-income-countries and more often undertreated: data from report-hf. European heart journal, 2020, 41(Supple2): null.

[27] Chugh S S, Roth G A, Gillum R F, et al. Global burden of atrial fibrillation in developed and developing nations. Global Heart, 2014, 9(1): 113–9.

[28] Gupta R, Wood D A. Primary prevention of ischaemic heart disease: populations, individuals, and health professionals. Lancet, 2019, 394(10199): 685–96.

[29] Addo J, Ayerbe L, Mohan K M, et al. Socioeconomic status and stroke: an updated review. Stroke, 2012, 43(4): 1186–91.

[30] Khaing W, Vallibhakara S A, Attia J, et al. Effects of education and income on cardiovascular outcomes: A systematic review and meta-analysis. European Journal of Preventive Cardiology, 2017, 24(10): 1032–42.

[31] Chambers J. Global and cross-country analysis of exposure of vulnerable populations to heatwaves from 1980 to 2018. Climatic change, 2020, 163(1): 539–58.

[32] Mensah G A, Roth G A, Sampson U K, et al. Mortality from cardiovascular diseases in sub-Saharan Africa, 1990-2013: a systematic analysis of data from the Global Burden of Disease Study 2013. Cardiovascular journal of Africa, 2015, 26(2 Suppl 1): S6–10.

[33] Upadhya B, Rocco M V, Pajewski N M, et al. Effect of Intensive Blood Pressure Reduction on Left Ventricular Mass, Structure, Function, and Fibrosis in the SPRINT-HEART. Hypertension, 2019, 74(2): 276–84.

[34] Sala L, Pontiroli A E. Role of obesity and hypertension in the incidence of atrial fibrillation, ischaemic heart disease and heart failure in patients with diabetes. Cardiovascular diabetology, 2021, 20(1): 162.

[35] Costas P, Garcia-Palmieri M R, Nazario E, et al. Relation of lipids, weight and physical activity to incidence of coronary heart disease: the Puerto Rico heart study. American journal of cardiology, 1978, 42(4): 653–8.

